# Temporal trends in online practices relating to autism and/or ADHD: a scoping review protocol

**DOI:** 10.1101/2024.05.01.24306633

**Authors:** Ruth Finn Leiser, Paul Flowers, Esperanza Miyake

## Abstract

**Background:** Increasing rates of neurodivergence diagnosis have received much attention in recent months. This is particularly true of autism and/or ADHD self-diagnoses precipitated by social media use. Mainstream media – at a minimum in English-speaking Western countries – has been quick to report on this, and it is clear that a social media-facilitated phenomenon is underway. To date, however, empirical evidence related to increased visibility of, and engagement with, autism and/or ADHD ‘content’ online over time is lacking.

**Objective:** To map temporal trends in online practices relating to neurodivergence – specifically confined here to autism and/or ADHD – within the published literature. Areas of interest include: how framing of the concept(s) change over time; the theories represented within research on this topic; whether the focal point of these online practices has changed over time.

**Inclusion criteria:** Published literature from any country and any time period after 1991, that relate to online activity involving what currently is often referred to as ‘neurodivergence’ – here specifically confined to autism and/or ADHD. ‘Online practices’ encompasses any aspect of online communication, information-seeking, support-seeking, awareness-raising, or associated practices that take place online – via search engines, chat rooms, forums, social media platforms. Studies looking at other conditions under the neurodivergence umbrella, and those pertaining to cyberbullying and internet addiction only will be excluded.

**Methods:** Following the Joanna Briggs Institute (JBI) methodology for scoping reviews, 4 databases (Web of Science, MEDLINE, CINAHL, and APA PsycInfo) will be searched. Inclusion criteria will be used to screen for, and select, appropriate studies. The JBI extraction tool will be adapted for this particular review, and the relevant data from included studies exported to this document. Both narrative accounts and figures of the data trends will be synthesised and presented.

## Introduction

Increasing rates of neurodivergence self-diagnosis – specifically related to Autism Spectrum Disorder (from herein referred to as ‘autism’), and Attention-Deficit/Hyperactivity Disorder (from herein referred to as ‘ADHD’) – have received much attention, and scrutiny, in recent years (de Broize et al., 2022; Gilmore et al., 2022). This is particularly true of self-diagnosis which has been precipitated by social media use, and the consumption of online content related to autism and/or ADHD (Russell et al., 2022; Graham, 2022). Both the National Institute for Health and Care Excellence (NICE, 2022) and the UK government’s Department of Health and Social Care (DHSC, 2022) have addressed the rise in adults seeking diagnosis of autism and/or ADHD, and an acknowledgement of historic under-diagnosis of autism in women led to significant funding into diagnostic pathways. There has also been extensive coverage of this trend across mainstream media (Graham, 2022). At the same time, individuals are taking to social media *en masse* to either discuss their own diagnostic journey or to seek guidance related to embarking upon it; many users assume an educator role – signposting others to what set them on their own path to diagnosis (Aragon-Guevara et al. (2023). It is clear that a social media-facilitated phenomenon is occurring, but to date, empirical experiential data on this is scarce (Botha & Cage, 2022).

Interest – and, indeed, academic research – into neurodivergence, however, is not new. Both autism and ADHD have long been speculated upon, theorised, and explored within academia and public health for decades (Volkmar & McPartland, 2014; Waltz, 2023; DiLorenzo et al., 2021). The enormity of the changes in our understanding of these neurotypes over time, however, cannot be overstated, and much of the research within this field has been rendered obsolete (den Houting et al., 2021; Dyck & Russell, 2020; Crompton, 2020) . From early theories of autism as ‘childhood schizophrenia’ and the result of ‘cold and detached’ parenting (Kanner, 1943; Bettelheim, 1967; Cleary et al., 2023), to studies concluding that food colourings cause ADHD (Boris & Mandel, 1994), numerous aspects of neurodivergence have only recently caught up with other advances in how we approach disability, especially within academia (Happé & Frith, 2020).

As awareness of autism and ADHD has grown, so too has academic – and public – perception changed. This is perhaps best perceived through the shift from language of pathology to that of neurodiversity (Craine, 2020; Singer, 1998; Bottini et al., 2023). Where, previously, autism was mostly theorised alongside learning disability – categorised by ‘severe’ vs ‘Asperger’s syndrome’ (the latter’s removal from the DSM in 2013 a testament to changes in the way autism is understood) (Pellicano & den Houting, 2022) – and conversations about ADHD mostly concerning an approximate moral panic about medicating ‘troublesome’ children (Swanson et al., 1995), this has changed in recent years. The paradigm shift from ‘disorder’ to ‘diversity’ has mainly surrounded neurodivergence in adults – something which empirical research had mostly neglected in previous years (Crane et al., 2018; Lipinski et al., 2022; Sonido et al., 2020).

Increased public perception of these neurotypes – and the associated experiences of living with them – has led to an influx of information online, ostensibly with the goal of educating, raising awareness, or increasing literacy around neurodivergence (Yeung et al., 2022). By virtue of the internet’s nature – a vast amalgamation of user-generated content with no built-in filtering mechanism for discerning validity or truth – this quest for signposting and educating has proven complicated, for a variety of reasons (Gilmore et al., 2022; Kang et al., 2017; Dewak, 2023). One issue is that of the veracity, trustworthiness, or accuracy of information being shared online; misinformation often the result of overzealous ‘awareness-raising’ that fails to separate individual experience from clinical diagnostic criteria (Aragon-Guevara et al., 2023). Similarly, as awareness of neurodivergence increases, so too do the scope of experiences which fit under the ‘neurodivergent’ umbrella (Eaton, 2023).

As diagnostic criteria are negotiated by both lay people and clinicians online, and clinical symptoms are intertwined with cultural identity and experiences, neurodivergence potentially becomes more relatable and easier to identify with. This increase in visibility and relatability – and arguably a ‘dilution’ of clinical diagnostic criteria – inevitably contributes to rising rates of diagnosis, which has attracted considerable levels of scrutiny, in relation to things like misinformation and pressure on diagnostic pathway waiting times (Gilmore et al., 2022). Despite this, there is a lack of empirical research on changing attitudes towards autism and ADHD diagnoses, and whether these reflect the rapid influx of people identifying as neurodivergent – encompassing either or both of these neurotypes. Similarly, exploration of academic research’s portrayal of autism and ADHD over time – from, for example, ‘childhood schizophrenia’ to ‘neurodiversity’ – could potentially provide some insight into the growing willingness for people to identify with conditions collected under the neurodivergent umbrella (Eaton, 2023).

One key component of temporal trends in neurodivergence awareness and acceptability, is undoubtedly the impact of the internet, and social media (Leadbitter et al., 2021). Society’s capacity for information exchange, and sharing of experience, has grown exponentially within the last few years (Ziebland & Wyke, 2012; Hage et al., 2020), and it can be assumed that this is reflected in societal attitudes towards autism and ADHD. The way people are using the internet in relation to autism and ADHD can be expected to have changed over the last 30 years – since the dawn of the World Wide Web – mirroring not only changes in attitudes towards neurodivergence, but also the technological advances of this time period and specific social media affordances that have emerged (Hassrick et al., 2021; (Ronzhyn et al., 2022). Mapping the academic literature on both neurodivergence and the online practices that are being conducted in relation to it, can provide insights into the cultural phenomenon that is taking place. The literature has, at time of writing, not yet been synthesised in such a way that maps these temporal trends, and therefore a review is required to chart the scope of the literature on these two key concepts, namely: 1) autism and ADHD as diagnoses and/or lived experiences, and 2) online practices relating to them. As there is a significant lack of empirical research on these concepts – both separately and combined – a scoping review is considered the best approach. This will enable the clarification of key definitions and concepts, and the identification of both extant literature and the presence of gaps.

A preliminary search of MEDLINE, the Cochrane Database of Systematic Reviews, and *JBI Evidence Synthesis* was conducted and no current or underway systematic reviews or scoping reviews on the topic were identified.

The objective of this scoping review is to systematically map the available literature on online practices relating to autism and/or ADHD, in order to identify gaps, quantify the areas of interest, and chart temporal trends. As the internet, and its varying online spaces, have changed over time, so too have the ways in which people are using the internet – and its various platforms – to engage with the concept of neurodivergence. Charting these changes over time – and technologies – can provide insights into how and why people are using the internet to engage with this topic, and also why the internet provides such a useful & appropriate space to explore issues of neurodivergence specifically.

### Review Questions

RQ1: Over time, how have autism/ADHD been framed within the published literature?

RQ2: Are there changes over time in the way that academic literature on autism/ADHD uses theory?

RQ3: Over time, how has the focal point of online practices^1^ relating to autism/ADHD changed within academic literature?

## Eligibility Criteria

The proposed scoping review will be conducted in accordance with the Joanna Briggs’ Institute (JBI) methodology for scoping reviews, using the ‘participant, concept, context’ (PCC) framework (Peters et al., 2020). See Appendix A for eligibility criteria table; full narrative account below.

### Participants

In contrast to the standard application of the JBI’s PCC framework, within this particular scoping review, participants are not defined for the purposes of inclusion/exclusion criteria. As the remit of this review is to identify, for example, technology, culture, identity, processes, and behaviours surrounding neurodivergence (specifically autism and ADHD), the focus is on the identified *concepts* (autism/ADHD and online practices), rather than participants. In other words, *who* is conducting this online activity – while of interest – does not form the basis of any inclusion/exclusion criteria. During data extraction, however, this information – i.e. *who* is primarily involved in this online activity – will be extracted from the selected studies and form part of the analysis. This will allow for trends and/or patterns to be identified within the literature on online practices relating to autism and/or ADHD, and will ensure that relevant studies are not precluded as a result of inclusion/exclusion criteria around specific participants and/or populations. It is expected that, within some literature, the participants/populations will be neurodivergent people themselves, and within other studies, will be stakeholders or those with a vested interest in neurodivergence, engaging in online practices relating to this. It is also anticipated that, in studies where the focus is on populations rather than concept (i.e. a sample of autistic people were recruited via the internet, but the study’s remit is not online practices relating to neurodivergence), this will be excluded based on ‘concept’ criteria.

### Concept

#### Inclusion

##### Neurodivergence

In this review, two specific facets of neurodivergence – autism and ADHD – are the focal point. This decision was made for a number of reasons. From a practical perspective, including all conditions/disorders/neurotypes included under the umbrella of neurodivergence – e.g. dyslexia, dyspraxia, developmental coordination disorder (DCD); dyscalculia; Tourette’s; auditory processing disorder (APD); sensory processing disorder (SPD); rejection sensitive dysphoria (RSD); Irlen Syndrome; cognitive functioning difficulties; executive dysfunction; dysgraphia; misophonia; obsessive compulsive disorder (OCD); synaesthesia; anxiety; trauma – was considered unwieldly to the point of being unworkable. For example, including all studies engaging with online activity relating to anxiety could populate a standalone review. In more conceptual terms, the extensive comorbidity between autism and ADHD – and, indeed, the extent to which this has engendered an increasingly-used moniker of ‘AuDHD’ – provided a solid basis for exploring these neurotypes both separately and as overlapping experiences. This clustering of autism and ADHD appears to be a unique phenomenon within the neurodivergence umbrella – no two other neurotypes are as closely linked together by both clinicians and lay people – nor has this comorbidity been comprehensively explained or researched. The emergence of ‘AuDHD’ as an established social phenomenon gives credence to its focus within this review.

##### Online practices

Studies engaged specifically with any internet-based practice that relates to neurodivergence will be included in this review. Web-based practices include, but are not limited to: viewing content on specific online applications (i.e. Facebook, Twitter, Instagram, TikTok); web browser searches (i.e. Googling information about autism); communicating via forums/chat rooms; health information-seeking searches/correspondence (i.e. searching online for signs/symptoms, engaging with those perceived to be knowledgeable on neurodivergence); joining online groups (i.e. those with a neurodivergence remit).

#### Exclusion

##### Neurodivergence

Any studies with a remit of neurodivergence that does not relate only to either autism, ADHD, or both. This includes studies on: dyslexia, dyspraxia, developmental coordination disorder (DCD); dyscalculia; Tourette’s; auditory processing disorder (APD); sensory processing disorder (SPD); rejection sensitive dysphoria (RSD); Irlen Syndrome; cognitive functioning difficulties; executive dysfunction; dysgraphia; misophonia; obsessive compulsive disorder (OCD); synaesthesia; anxiety; trauma.

##### Online practices

Where online means have been used to recruit participants or host data collection (i.e. web-based video chat, or online surveys) within a study that involves neurodivergent participants or neurodiverse samples – but the research itself does not relate to online activity regarding neurodivergence – these will not be included.

At the intersection of both aspects of this review’s concept, a consistent result within preliminary searches were studies relating to cyberbullying. This appears to be a major issue within research on neurodivergence experiences online. It was decided, however, that ‘cyberbullying’ would not be included within the remit of this review. This is the result of extensive discussion, and theorising, within the wider team. Eventually, the concept of this review was conceptualised as relating to people actively seeking out content, support, information, advice, diagnostic criteria online. Regardless of the person conducting this online activity – as previously stated, ‘Participant’ is not defined here – the online activity was still being conducted by the/a person actively seeking out neurodivergent-adjacent online content/correspondence. Cyberbullying, on the other hand, is conceptualised as being conducted by a separate and distinct ‘participant’ altogether, with extremely distinct motivation(s) and for unrelated reasons. Therefore, literature that pertains to cyberbullying of neurodivergent people will be excluded during the screening process.

Similarly, where a study design uses a web-based chat room assessment/task to observe/test a specific phenomenon, but this forms the entirety of ‘online activity’ within the study, this will be excluded. This is due to the online activity in this study merely acting as a conduit for the research’s true aim and/or objective, rather than the research pertaining to online activity itself. Where studies develop chat room-based interventions, however, which are intended for people to continue using after the data collection, and not just as a tool within the study itself, this will be included. Similarly, any study with the remit of observing or reporting, for example, organic chat room use by a neurodivergent population, will be included.

A final, yet notable, area for selective inclusion/exclusion is internet addiction, web-based gaming addiction, or ‘problematic internet use’. While there does seem to be a particular phenomenon occurring with internet addiction and neurodivergence – and of particular interest is the potential cultural differences that may be found – i.e. framing socialising online as useful vs unproductive/problematic – this is beyond the scope of this review. The quantity of papers relating to internet addiction and neurodivergent populations is also unmanageable in terms of screening and data extraction within the available timeframe (e.g. from one of the review’s selected databases, inclusion of these concepts produced approximately 900 additional papers). Within these papers, however, it is acknowledged that a very small number may relate specifically to either the neurodivergent diagnostic journey or the lived experience of social interaction amongst neurodivergent people, facilitated through ‘problematic’ internet use. Where these specific situations occur, studies will be included within the review. If a study is simply a descriptive account of population parameters or prevalence of interaction addiction, it will be excluded during the title & abstract screening stage.

### Context

#### Inclusion

##### Location

Given that this review’s remit does not preclude people whose self-diagnosis of either autism or ADHD was the basis for formal diagnosis – and therefore can include those whose self-diagnosis simply informed their own sense of identity – there was no requirement for limiting results by geographical location. Similarly, this increase in awareness – and therefore diagnosis – is considered a global phenomenon; the crucial role of the internet means that studies from all countries will be included – however, results will be limited to English-language only.

##### Timescale

Due to the nature of this review’s objectives (i.e. temporal charting of online practices/trends), the timescale imposed upon study inclusion will be constrained by the dawn of the World Wide Web. Given that the Web was not made public until 1991, this forms an objective start point to the review’s scope.

#### Exclusion

##### Timescale

Literature published prior to 1991, here considered the ‘dawn’ of the World Wide Web.

### Types of sources

#### Inclusion

Owing to the public discourse that has surrounded autism and ADHD for decades, this review lends itself to an inclusive approach to sources – for example, inclusion of letters (i.e. research letters, letters to the editor) & opinion pieces. As a result, there will be no parameters set for inclusion within published literature.

#### Exclusion

For the purposes of conducting a timely review, dissertations/doctoral theses will be excluded. This is to facilitate workability of screening within an appropriate timeframe.

## Method

The proposed scoping review will be conducted in accordance with the JBI methodology for scoping reviews (Peters et al., 2020).

### Search strategy

The search strategy will aim to locate published studies. An initial limited search of Web of Science, and APA PsycINFO was conducted in December 2023, in order to identify an initial set of articles related to the topic.

The PCC framework will be used to help structure the search – most notably with inclusion/exclusion criteria – but in terms of defining actual search terms, the ‘participant’ and ‘context’ criteria are considered less relevant. ‘Participant’, as mentioned above, has been removed altogether from the inclusion/exclusion criteria, and therefore does not play a particularly central role in structuring the search strategy – although it will underpin the data extraction process. ‘Context’, too, does not feature prominently in the search terms, but will factor in during the search itself, and the subsequent screening process – i.e. search limiters will include ‘post-1991’, and ‘English only’.

A preliminary search of APA PsycInfo and MEDLINE was conducted, using the keywords ‘online’ and ‘autism OR ADHD OR neurodivergent’, to find initial relevant papers. Subsequently, analysis of the text words within the title and abstract of the papers, and additionally the index terms used to describe the articles, was conducted. This provided the basis for a full search strategy to be developed – including all appropriate search terms for the review’s defined concept(s), as well as search limiters like date range and language. This search strategy was then adapted for each included database – e.g. where differences exist between field entry labels. Included databases were as follows: APA PsycInfo, MEDLINE, CINAHL, and Web of Science. See Appendix B for full search example.

Gray literature has been excluded from this review. The main reason for this relates to the comparative poor quality of sources due to lack of peer review. Increasing rates of neurodivergent diagnosis – both formal and self-reported – have been the subject of much scrutiny and controversy globally, and inclusion of literature which has not been peer-reviewed is unlikely to produce suitably coherent and relevant contributions.

The reference list of all included sources of evidence will be screened for additional studies (Jenkins, 2004).

Included study language will be limited to English only. This is due to lack of readily available translation capacity within the supervisory team. Studies published since 1991 will be included, as this is the year the World Wide Web was made public, and therefore is considered the appropriate start point for online activity by the public.

### Study/Source of evidence selection

Following the search, all identified citations will be collated and uploaded into Rayyan (Ouzzani et al., 2016) and duplicates removed.

Titles and abstracts will then be screened by RL (100%) and GE (10%) for assessment against the inclusion criteria for the review.

Following screening, the full text of selected citations will be assessed in detail against the inclusion criteria by RL and GE. Reasons for exclusion of sources of evidence at full text that do not meet the inclusion criteria will be recorded and reported in the scoping review. Any disagreements that arise between the reviewers at each stage of the selection process will be resolved through discussion, or with an additional reviewer (JMcL/PF). The results of the search and the study inclusion process will be reported in full in the final scoping review and presented in a Preferred Reporting Items for Systematic Reviews and Meta-analyses extension for scoping review (PRISMA-ScR) flow diagram (Tricco et al., 2018).

### Data Extraction

Data will be extracted from papers included in the scoping review using a data extraction tool adapted from the JBI Manual for Evidence charting table for data extraction synthesis (Peters et al., 2020). The data extraction process will be shaped by the PCC framework and will include specific details about the participants, concept, context, study methods, and key findings relevant to the review questions.

A draft extraction form is provided (Appendix C). The draft data extraction tool will be modified and revised as necessary during the process of extracting data from each included evidence source. Modifications will be detailed in the scoping review. Any disagreements that arise between the reviewers will be resolved through discussion, or with an additional reviewer/s. If appropriate, authors of papers will be contacted to request missing or additional data, where required.

### Data Analysis and Presentation

For all research questions, given that temporal trends within published literature are being explored – in addition to tabular and narrative summaries, the data will also be presented within a graph or figure that will visually convey any relevant patterns over time.

## Data Availability

All data produced in the present study are available upon reasonable request to the authors

## Appendices

### Appendix A

*Scoping review eligibility criteria table*

|  |  |
| --- | --- |
| **Participants** |  |
| *Inclusion* | N/A – see *Participants* section above, pg6 |
| *Exclusion* | N/A – see *Participants* section above, pg6 |
| **Concept** |  |
| *Inclusion* | **Neurodivergence**<br>Autism; ADHD/ADD; “AuDHD”<br>**Online practices**<br>Any internet-based practice that relates to the specified neurotypes above. Web-based practices include viewing content on specific online applications; web browser searches; communicating via forums/chat rooms; health information-seeking searches/correspondence; joining online groups etc. This includes, but is not limited to: Facebook; Twitter; TikTok; Instagram; YouTube; Reddit; social media; social platforms; social networking sites; internet; web; chat rooms; forums |
| *Exclusion* | **Neurodivergence**<br>Dyslexia; Dyspraxia; Development Coordination Disorder (DCD); Dyscalculia; Tourette’s Syndrome; Auditory Processing Disorder (APD); Sensory Processing Disorder; Rejection Sensitive Dysphoria (RSD); Irlen Syndrome; Cognitive functioning difficulties; Executive dysfunction; Dysgraphia; Misophonia; Obsessive Compulsive Disorder (OCD); Synaesthesia; Anxiety; Trauma<br>**Online practices**<br>Online practices which do not specifically relate to neurodivergence, but were used as a method of i.e. recruitment or survey dissemination within a study that involved neurodiverse samples or neurodivergent participants |
| **Context** |  |
| *Inclusion* | **Geographic location**<br>Any; English-language only<br>**Timescale**<br>Post-1991 |
| *Exclusion* | Literature published prior to 1991 |
| **Types of sources of evidence** |  |
| *Inclusion* | Any source of published evidence, with the exception of dissertations/theses |
| *Exclusion* | Dissertations/theses |

### Appendix B: Search strategy/example

*Example search strategy, including search terms, conducted on APA PsycInfo (EBSCO)*

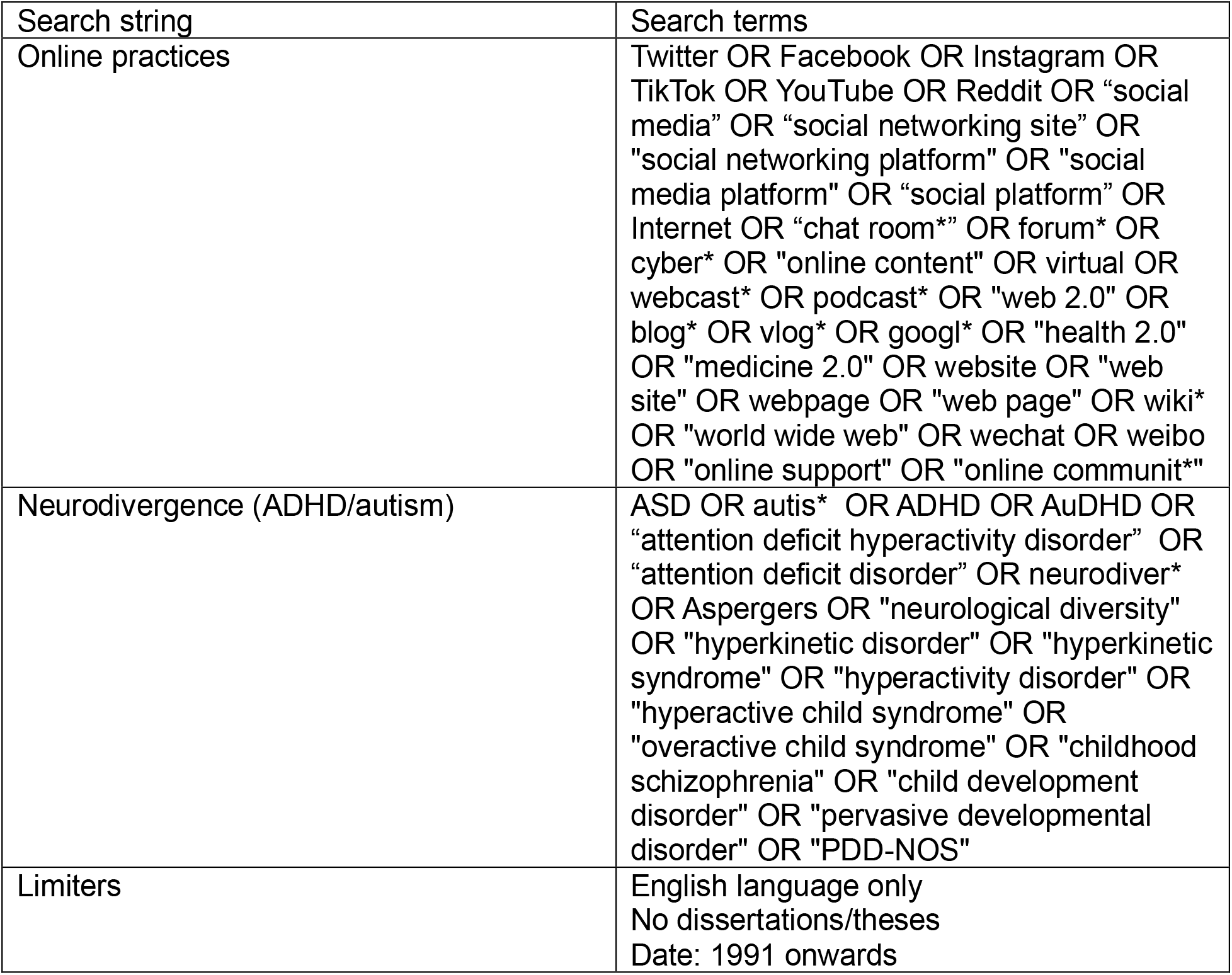

### Appendix C: Draft data extraction form

*Table constructed from data extraction Excel spreadsheet, including all relevant information to be extracted from included papers during scoping review*

| **Category** | **Data to be extracted** |
| --- | --- |
| Publication details | Author<br>Year<br>Aim/Objective<br>Research Question |
| Study details | Field/area of research<br>Theories/frameworks used<br>Research design<br>Data collection<br>Analysis<br>Sample size<br>Recruitment strategy |
| Participant | Participant(s) of the study<br>Participant diagnosis status – i.e. autism, ADHD, ‘AuDHD’<br>Diagnosis type – i.e. formal diagnosis, self-diagnosis, self-diagnosis followed by formal diagnosis etc<br>Stakeholder status – i.e. participant(s) are not neurodivergent, but are stakeholders of the neurodivergent community<br>Age of participant(s) – either specific age group, age range, or not specified<br>Support needs of participants – i.e. studies which sample only high support needs individuals, versus studies that involve low support needs individuals, or not specified |
| Concept | Neurotype investigated by study – i.e. autism, ADHD, ‘AuDHD’<br>Framing of neurotype – i.e. diversity, disability, impairment etc<br>Online platform/tool investigated by study – i.e. does the study specifically explore online practices on Facebook, or web searches, or is this not specified<br>Main reason for use – i.e. what facet of online practices does the study explore – communicating, support-seeking, information-seeking etc<br>Time spent on the online platform/tool |
| Context | Location – i.e. where data collection took place<br>Year of data collection |
| Other | Type of evidence – i.e. what format does the publication take<br>Findings/conclusion of the study |

## Footnotes

1 Online practices here relates specifically to conducting searches, communication, information-seeking etc directly related to autism and/or ADHD – and therefore excludes secondary online activity like cyberbullying and internet addiction.

